# Transdiagnostic Approach in Cerebral Palsy

**DOI:** 10.64898/2026.04.27.26351832

**Authors:** Philip E. Gates, Charlotte A. Chun, Louise C. Bonneau, Demiana A. Soliman

## Abstract

**OBJECTIVES:** Demonstrate correlations of clinic-based measures of International Classification of Functioning, Disability and Health (ICF) Body Structure and Function, capacity and performance with a school-based performance measure in children with Cerebral Palsy (CP) using a transdiagnostic approach.

**METHODS:** 102 ambulatory children with CP underwent assessment of Gross Motor Function Classification System (GMFCS), Gross Motor Function Measure (GMFM), Pediatric Quality of Life Inventory Generic Core Scales (PedsQL), 3-Dimensional Gait Analysis, Gillette Functional Assessment Questionnaire (GFAQ), and Pediatric Outcomes Data Collection Instrument (PODCI) done in clinics, compared with School Function Assessment (SFA) done in schools. Here we report on SFA correlations. For this paper, Spearman’s correlations were calculated.

**RESULTS:** All measures showed some significant correlations with the SFA; greatest number of moderate to strong correlations were with PODCI, including PODCI comorbidities scales. PODCI performance questionnaire was correlated with all SFA scales. PODCI, as a performance measure, is broader, more holistic, than the capacity and BSF measures. Findings are demonstrative of a focus on the ICF approach, indicating separate domains of function and well-being, reflective of the transdiagnostic approach.

**CONCLUSIONS:** The transdiagnostic approach, looking at a broader picture than simply diagnosis, thus paralleling concepts presented in the ICF, is beneficial in assessing functioning and well-being in children with CP.

## INTRODUCTION

### Transdiagnostic Approach

In this century, medicine has two driving forces. One is a strong push toward “precision medicine,” which focuses increasingly on genomics and machine learning. The second is a World Health Organization (WHO) effort called the International Classification of Functioning, Disability and Health (ICF), which considers health and functioning with a broader lens. [1] This article addresses functioning and well-being based upon the ICF approach, “measuring functioning in society, no matter what the reason for one’s impairments.” [1] We argue for the value of assessing “functioning, disability, and health” across diagnoses, or the *transdiagnostic* approach, which 1) facilitates assessment of functioning, crucial for treatment planning and outcome evaluation, 2) allows identification of shared mechanisms spanning multiple conditions, and 3) represents individuals’ lived experiences more realistically. The term *transdiagnostic* has appeared in almost 5000 peer-reviewed articles in mental health since it first appeared on PubMed in 2007. Though this terminology has not been used in physical health articles, we show the transdiagnostic approach is relevant in physical health management.

### Cerebral Palsy

We approach this topic by way of a multicenter study completed in 2005 and partially reported in two articles [2,3]. Here, we share more findings from the study and focus on its foundation in the ICF. The study was of school-aged children with Cerebral Palsy (CP), correlating a validated measure of performance in school (School Function Assessment, SFA [4]) with measures of body function and structure, capacity, performance, and health-related quality of life (HRQOL) in clinic.

The Centers for Disease Control (CDC) defines CP as “a group of disorders that affect a person’s ability to move and maintain balance and posture … caused by abnormal brain development or damage to the developing brain that affects a person’s ability to control his or her muscles.” [5] People with CP present differently: some merely walk with an unusual gait while others may not walk at all and therefore require constant care. “CP does not get worse over time, though the exact symptoms can change over a person’s lifetime. All people with CP have problems with movement and posture, which indicates musculoskeletal impact. Many also have related conditions such as intellectual disability; seizures; problems with vision, hearing, or speech; changes in the spine (such as scoliosis); or joint problems (such as contractures).” (5) Therefore, people with CP often require care for more than CP: they benefit from holistic care, or the transdiagnostic approach. (“…problems with movement and posture” indicates having “musculoskeletal impact”)

### International Classification of Functioning, Disability and Health

This ICF diagram illustrates how “disability and functioning are viewed as outcomes of interactions between health conditions (diseases, disorders and injuries) and contextual factors,” environmental or personal [1]. ICF (Figure 1) explains that “among contextual factors are external environmental factors (for example, social attitudes, architectural characteristics, legal and social structures, as well as climate, terrain and so forth); and internal personal factors, which include gender, age, coping styles, social background, education, profession, past and current experience, overall behavior pattern, character and other factors that influence how disability is experienced by the individual.” [1] If body functions and structure, activity, and participation are factors of human functioning, then disability is dysfunction in one or more of those functions. Two qualifiers explain the severity of dysfunction:

**Figure 1.**
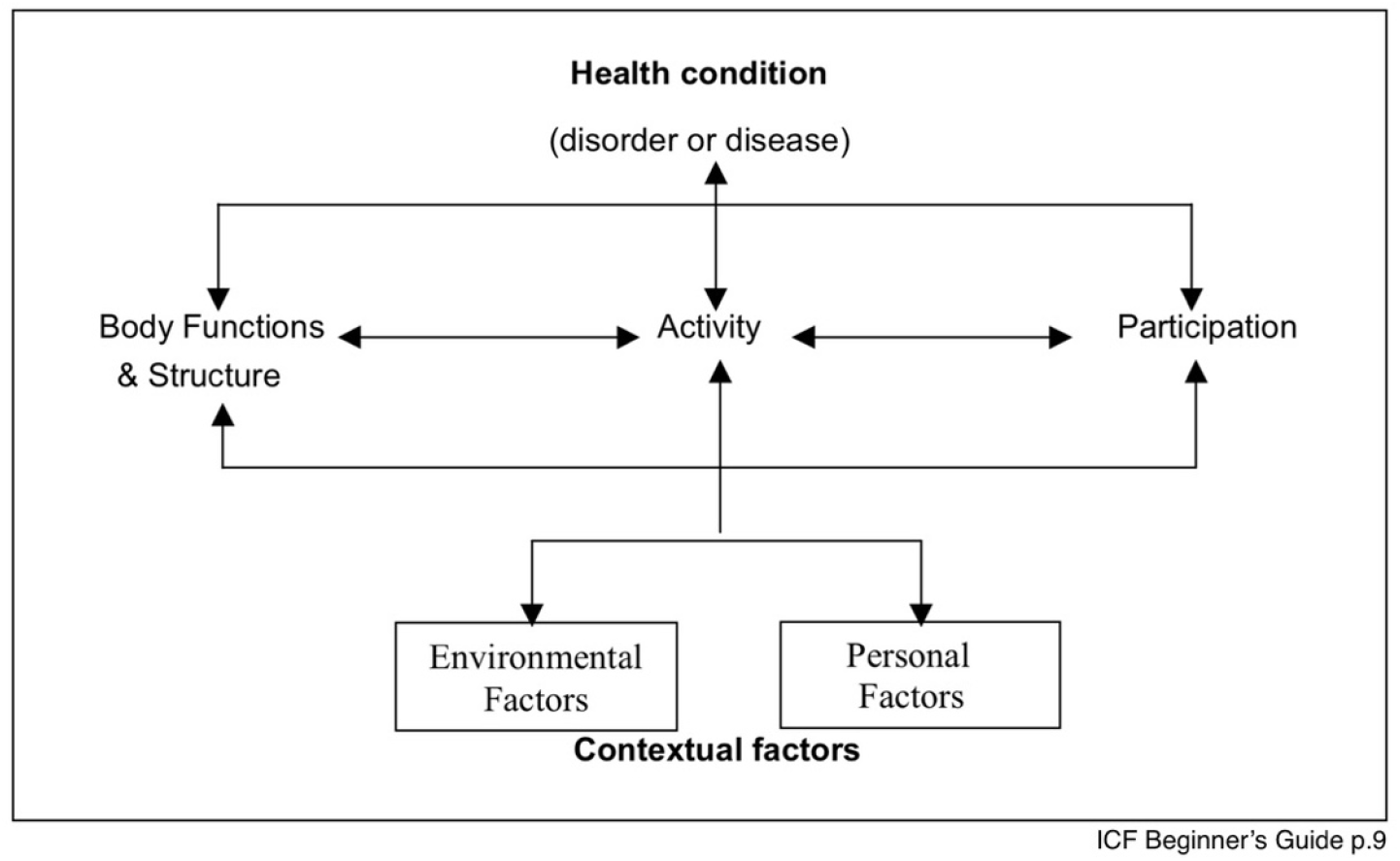
ICF Diagram illustrating outcomes of Contextual factors and health conditions interactions. Interactivity of domains of International Classification of Functioning, Disability and Health.

- “The performance qualifier describes what an individual does in his or her current environment.” [1] This includes a broad understanding of a person’s general social context as well as a narrow understanding of that person’s individual experience, including any required assistance.
- “The capacity qualifier describes an individual’s ability to execute a task or an action.” [1] This includes a generous understanding of a person’s ability to function in a certain place and time without assistance. [1]

This multicenter study, first published in a limited fashion in 2008, assessed correlations between performance in the community and measures of body structures and function (BSF), capacity, performance and HRQOL in the clinic for children with CP. We report more extensive results here to establish the relevance of this study to transdiagnostic terminology, as supported by two related articles in process of submission for publication.

## METHODS

Based upon the concepts in the ICF Beginner’s Guide, we undertook a multicenter study, completed in 2005, partially reported elsewhere [2,3]. The study was approved by Institutional Review Board (IRB) review at each of the three participating centers. We used a validated measure of school performance (SFA) against which we compared clinic-based measures. Correlations indicated the tested clinic measures reflect school functioning reported by those working with the children daily in school. “SFA facilitates collaborative program planning for students with various disabling conditions.” [4] Measures assessed include GMFM (assesses capacity) [6], GFAQ (more capacity questions than performance) [7], PedsQL (HRQOL) [8], 3-Dimensional Gait Analysis (measures gait/body structure and function in idealized environment of the Motion Analysis Center), and PODCI (primarily focuses on performance, with some capacity and contextual questions) with comorbidities scales. PDF file of the PODCI form is attached as supplemental Data [9]. PODCI and PedsQL are more holistic. Severity of CP was rated by clinicians using the Gross Motor Function Classification System Expanded and Revised (GMFCS Level Determination - https://www.canchild.ca/system/tenon/assets/attachments/000/000/058/original/GMFCS-ER_English.pdf) [10] here we simply designate as GMFCS).

The multicenter study had 102 children in the final report. Of those, eight had no reported PODCI completed. Of the remaining 94 children, 61 required no assistive device (GMFCS I-II) and 33 required an assistive mobility device (GMFCS III-IV). PODCI proxy report in clinic demonstrated ecological validity with SFA, establishing PODCI as a performance measure. PODCI also includes measures of well-being (HRQOL), demographics and comorbidities. Gates et al 2008 reported on correlations between PODCI and SFA, especially focusing on details of individual questions. “The strongest indicators of a child’s overall performance in school appeared to be three PODCI items: PODCI28, ‘Is it easy or hard for your child to button buttons?’; PODCI48, ‘How often does your child need help from another person for walking or climbing?’; and PODCI56, ‘How often does your child need help from another person for sitting or standing?’” They also reported significant correlations with contextual factors [2].

Rabinovich et al reported correlations of SFA with GMFCS. Spearman’s rank correlation comparing SFA to GMFCS showed significant correlation between the composite SFA criterion score and GMFCS class (r = -0.847, p < 0.02) (3). Here we share all correlations of assessed measures and focus on the broad correlation of PODCI with SFA, highlighting its more holistic results.

### Statistical Methods

Analysis was performed using SPSS version 29.0.2.0. Because most variables were non-normally distributed and residuals of their regression with SFA variables deviated from normality (one-sample Kolmogorov-Smirnov tests *p* <.05, indicating significant non-normality), associations were examined with Spearman rank-order correlations, a non-parametric test appropriate for variables with these distortions.

## RESULTS

There were more males than females (approx. 59% to 41%). Most had government or private insurance or both; approximately 15% were uninsured. Estimated median income based upon zip code ranged from $21,000 to $126,000. A limited number of children had GMFM assessment (33) and PedsQL [24] compared to other assessments (86-101). Table 1 includes sample sizes for each measure; missing data varied across measures.

**Table 1.** Patient Demographics.

|  | **N** | **Range** | **Mean (SD)** |
| --- | --- | --- | --- |
| Age | 101 | 5 – 18 | 11.76 (3.24) |
| Gait Velocity | 47 | 21 – 150 | 65.58 (25.79) |
| Gross Motor Function Classification System | 95 | 1 – 4 | 2.22 ( 0.70) |
| Gillette Functional Assessment Questionnaire Total | 84 | 8 – 77 | 45.45 (13.60) |
| Gross Motor Function Measure (GMFM) – Total Score | 33 | 16 – 96 | 61.67 (21.15) |
| GMFM – Section D: Standing Score | 33 | 9 – 36 | 27.21 (5.90) |
| GMFM – Section E: Walking, Running, and Jumping Score | 33 | 3 – 60 | 34.45 (16.01) |
| Pediatrics Outcomes Data Collection Instrument (PODCI) Comorbidities | 86 | 0 – 14 | 4.76 (3.19) |
| PODCI Global | 92 | 38 – 100 | 73.29 (12.68) |
| PODCI Upper Extremity | 95 | 33 – 100 | 80.56 (15.08) |
| PODCI Transfers and Basic Mobility | 92 | 26 – 100 | 80.26 (15.21) |
| PODCI Sports and Physical Function | 95 | 6 – 100 | 52.93 (20.42) |
| PODCI Comfort | 95 | 26 – 100 | 80.45 (20.36) |
| PODCI Happiness | 94 | 20 – 100 | 77.89 (20.57) |
| Pediatric Quality of Life Inventory (PedsQL) – Total Score | 24 | 0 – 22 | 9.25 (6.33) |
| PedsQL – Physical Functioning | 24 | 0 – 9 | 3.21 (2.69) |
| PedsQL – Social Functioning | 24 | 0 – 8 | 2.71 (2.37) |
| PedsQL – School Functioning | 24 | 0 – 7 | 1.83 (2.22) |
| PedsQL – Emotional Functioning | 24 | 0 – 4 | 1.50 (1.59) |
| School Function Assessment (SFA) |  |  |  |
| SFA Travel | 99 | 25 – 76 | 67.28 (9.76) |
| SFA Maintaining and Changing Position | 101 | 13 – 48 | 42.58 (7.13) |
| SFA Recreational Movement | 95 | 11 – 44 | 28.91 (10.78) |
| SFA Manipulation With Movement | 101 | 20 – 64 | 53.13 (10.54) |
| SFA Using Materials | 101 | 32 – 100 | 87.63 (15.45) |
| SFA Setup and Cleanup | 100 | 24 – 64 | 59.12 (7.51) |
| SFA Eating and Drinking | 101 | 32 – 64 | 54.62 (4.17) |
| SFA Hygiene | 101 | 22 – 70 | 57.36 (6.43) |
| SFA Clothing Management | 97 | 26 – 68 | 58.80 (12.71) |
| SFA Up & Down Stairs | 68 | 4 – 24 | 15.82 (6.42) |
| SFA Written Work | 99 | 12 – 53 | 38.62 (9.32) |
| SFA Computer & Equipment Use | 72 | 10 – 36 | 27.44 (5.39) |
| SFA Functional Communication | 100 | 25 – 54 | 49.75 (4.31) |
| SFA Memory and Understanding | 100 | 19 – 40 | 37.88 (3.67) |
| SFA Following Social Conventions | 100 | 28 – 48 | 45.37 (4.24) |
| SFA Compliance with Adult Directives and School Rules | 99 | 34 – 70 | 56.64 (6.05) |
| SFA Task Behavior/Completion | 100 | 31 – 76 | 67.40 (10.70) |
| SFA Positive Interaction | 100 | 29 – 80 | 71.72 (11.69) |
| SFA Behavior Regulation | 98 | 13 – 48 | 43.22 (7.14) |
| SFA Personal Care Awareness | 98 | 14 – 40 | 37.61 (4.81) |
| SFA Safety | 98 | 10 – 40 | 37.32 (5.87) |

Table 2 shows that overall, measures in the clinic demonstrated varying correlations with SFA. Capacity measures (GMFM and GFAQ) and body structure and function measures (Gait velocity) do not show as many medium or large effect sizes, correlating with school performance, substantiating ICF taxonomy. Correlations of scales are noted as p<0.01 with ** and p<0.05 with *. However, more telling are the effect sizes (11). PODCI scales with greatest correlation with SFA scales are Comorbidities, Upper Extremity, Transfers and Basic Mobility, and Sports and Physical Function.

**Table 2:**
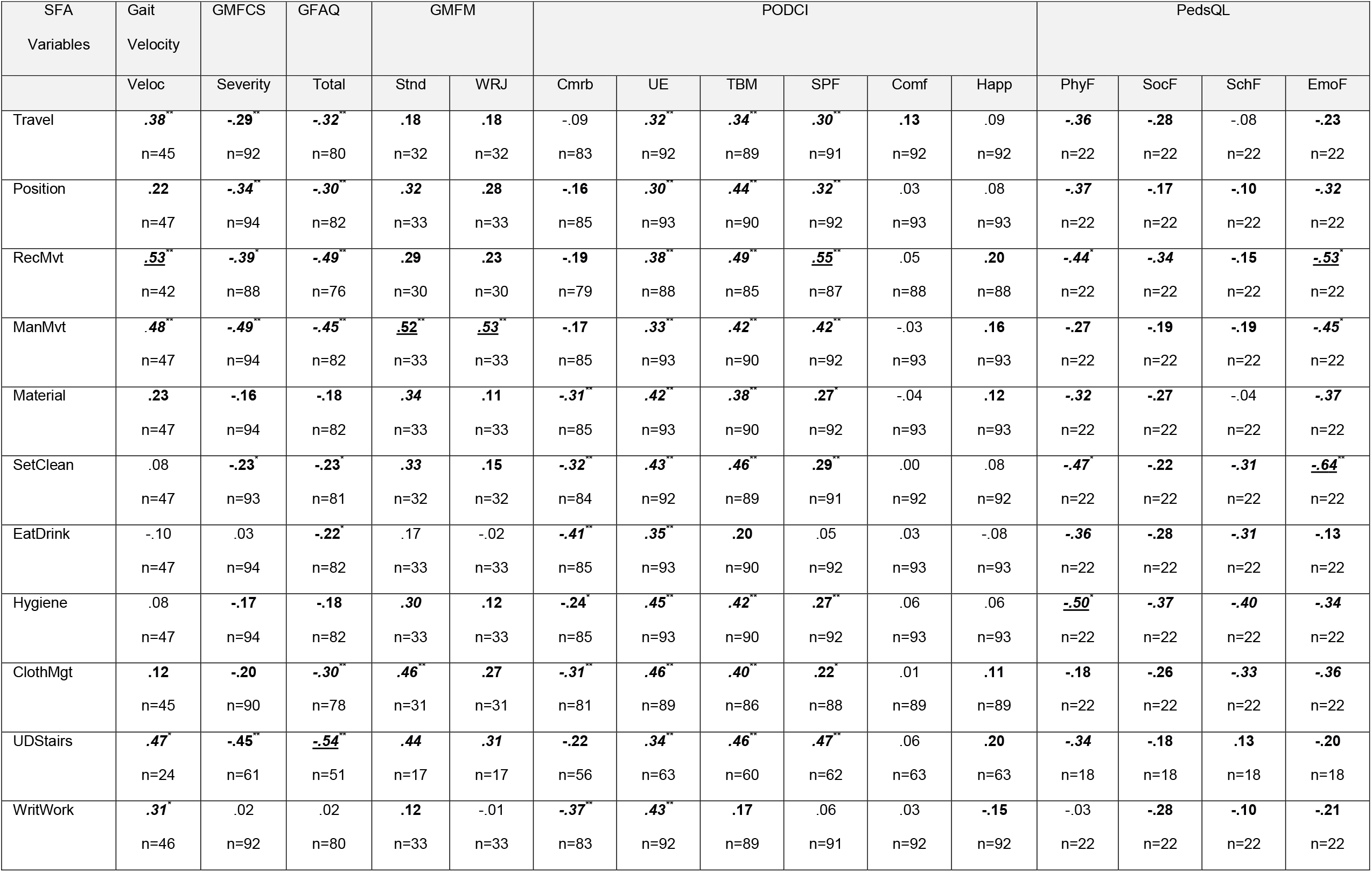
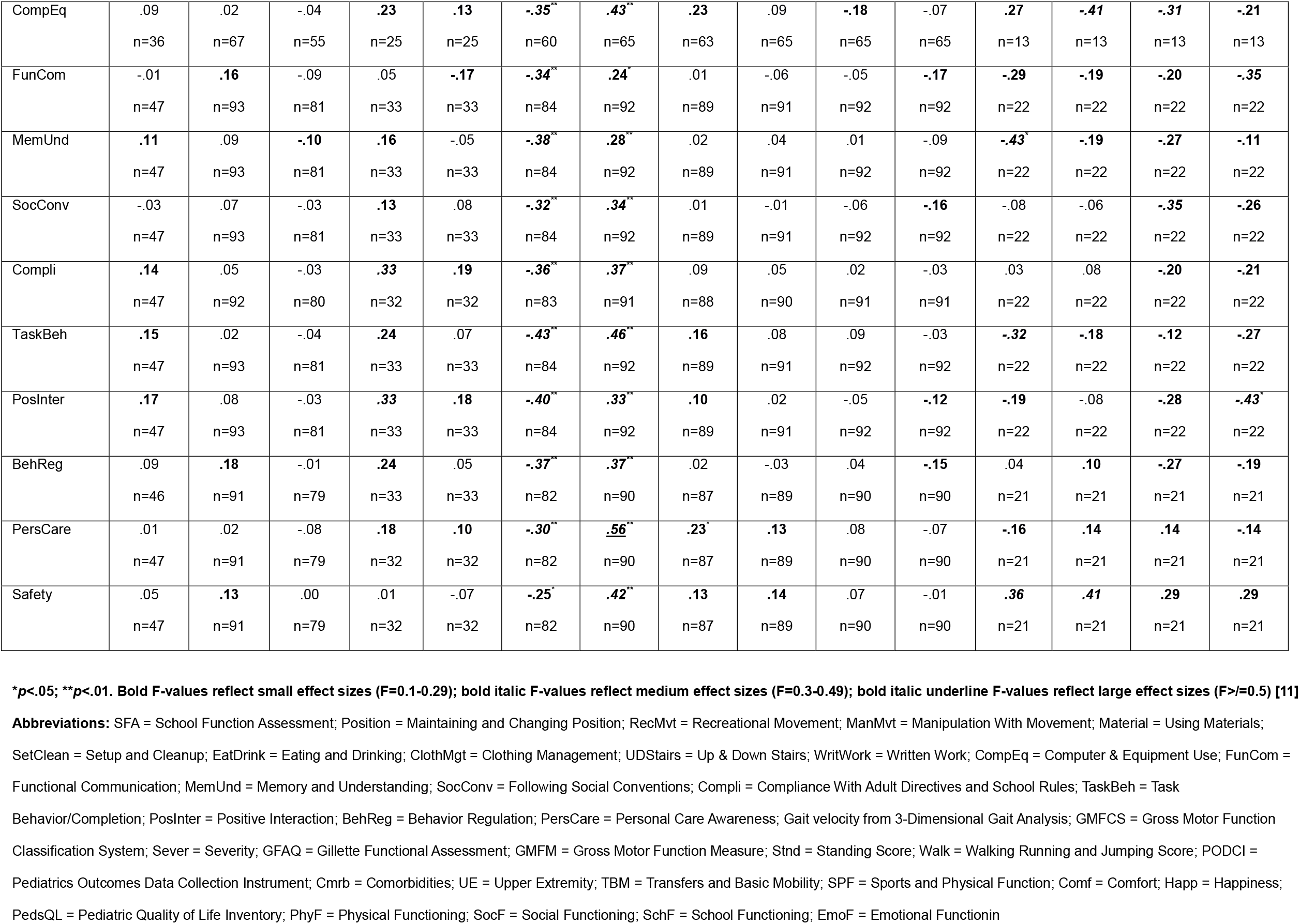
Correlations with interpretations between SFA Scales and measures of body structure and function (Gait velocity), capacity (GMFM and GFAQ), performance (PODCI), and HRQOL (PedsQL and PODCI) Spearman’s rank-order correlations between SFA variables and Clinical Variables (F-values)

In general, the HRQOL (PedsQL), performance (PODCI, GMFCS), capacity (GFAQ, GMFM), and functional measures (Gait velocity) all showed some significant correlations with SFA scales at the level of a small-to-medium effect size.

In general, physical subscales of SFA showed stronger associations with HRQOL (PedsQL), performance (PODCI, GMFCS), capacity (GFAQ, GMFM), and functional (Gait velocity) measures than cognitive-behavioral scales of SFA.

Overall, the PODCI showed moderate correlations with the SFA scales. There was variability across PODCI scales: strongest correlations (medium effect size) were found between the PODCI scales assessing personal factors (Comorbidities - Cmrb) and fine/gross motor function (Upper Extremity (UE), and Transfers and Basic Mobility (TBM) and the SFA versus relatively weaker correlations (negligible to small effect sizes) between the PODCI HRQOL scales (Comfort, Happiness - Happ).

Overall, PedsQL showed small-to-moderate correlations with SFA scales. There was some variability across subscales with Emotional Functioning scale showing the strongest correlations across the board.

## Discussion

Some of the detail of the multicenter study with PODCI and SFA was reported in Gates et al, 2008 [2] and GMFCS and SFA in Rabinovich [3]. In general, quality of life, performance, capacity, and functional measures all showed small to moderate associations with school performance, demonstrating the ecological validity of these measures. This means that parents and clinicians provide valid ratings of the child’s functioning, matching what school personnel witness day to day. Gates et al, 2008 noted, “The results of this study provide strong evidence that clinicians can accurately assess a child’s participation in the community by collecting health-related quality of life data from PODCI in the clinic setting.” [2] In addition, the ecological validity of proxy report of PODCI is confirmed.

Elsewhere GMFM [6], GFAQ [7], Walking speed [11], though in young adults) and PedsQL [8] have been used successfully in CP. One thing this means is that for routine evaluations of school performance it is not necessary to enlist the assistance of school personnel in working through the time-intensive 286-question SFA.

Overall, PODCI showed moderate correlations with SFA. The findings are consistent with the previous report [2], here showing more extensive results. Most significant is the reinforcement that collecting comorbidity data is important for this population (also noted in [14]), consistent with the “transdiagnostic theme.” Moderate level correlations are seen in Upper Extremity and Physical Function and Transfers and Basic Mobility scales with SFA scales assessing physical performance, and Upper Extremity and Physical Function even with cognitive-behavioral scales of SFA.

Overall, PedsQL showed small-to-moderate correlations with SFA, but the Emotional Functioning scale showed the strongest correlations across the board, suggesting that clinicians or researchers specifically interested in HRQOL/Emotional functioning issues might use PedsQL.

Our purpose is to introduce novel terminology for the medical approach to managing children with movement disorders that parallels an approach widely used in mental health, the transdiagnostic approach. This approach “cuts across traditional diagnostic boundaries or, more radically, sets them aside altogether.” [6] Our research supports this application to medical treatment of children with movement disorders because it allows a multi-faceted approach to care, helping clinicians gain a holistic understanding of patients and helping children function well in community. The transdiagnostic approach makes whole-person care the standard of care, broadly assessing health conditions as well as contextual factors when determining treatment.

Because humans are complex organisms, we almost always benefit from a transdiagnostic approach, but persons with disabilities require it. Transdiagnostic techniques are well-validated: the term simply describes a method that functional clinicians have always used, focusing on symptoms that are present in multiple disorders to offer treatments relevant to a patient’s practical, real-world experience. The term is common in psychiatric literature [13] and has been used in some childhood-onset disabilities like autism (274 references PubMed 02082024) and ADHD (251 references PubMed 02082024).

Children with movement disorders need treatment that considers the relationships between symptoms shared by different diseases. No one clinical approach can provide an accurate understanding of these patients’ conditions or provide effective treatment. We need the transdiagnostic approach because it focuses on concepts and allows us to use dimensional models to examine constructs along a spectrum instead of a yes/no diagnosis. Diagnoses do not always indicate certain functional outcomes, and complex conditions require especially broad consideration of symptoms. The transdiagnostic approach is a whole-child approach that bridges this gap.

The three centers involved in the multicenter study reported here are regional pediatric orthopaedic referral centers covering large geographical areas, serving children by means of clinic services, rehabilitation services, orthotics and prosthetics, 3-D motion analysis, radiographic screening for hip dysplasia and scoliosis, and surgical services. Specialized clinics were clustered so that children and families could have “community,” interacting with others facing similar challenges. Children and families with CP and other childhood-onset disabilities benefit from peer interactions to combat physical and emotional isolation, which is especially common in our population. Because a large percentage of our catchment areas is rural, the children were often their Primary Care Physician’s only patient with CP. Thus, many received supportive care at our facilities and learned that they were not “all alone.”

The mental health benefits of this sense of community support a patient’s physical treatment; both contribute to total functioning. So too, children with movement disorders report finding camp experiences vital to address isolation and prepare for adult independence. Louise C. Bonneau, LCSW, a former clinic patient with cerebral palsy, spastic quadriplegia, says, “We need places like Shriners and Lions Camp where for a little while the world is made for us, where we can interact with others who ‘get it.’” This is not a need that patients outgrow. Transition to adulthood is always challenging for children with movement disorders because so few services are available for adults with CP. A transdiagnostic approach helps clinicians view various aspects of functioning together and compare effects across diagnoses.

One functional effect of disability that most clinical tools fail to assess is what ICF identifies as an individual’s experience of disability. One common challenge is ableism, defined by Neumeier and Brown [15] as the idea that only some people’s brains or bodies are healthy, whole, functional, and valuable in society while others are broken, defective, inferior, and unworthy. Ableism values people who seem “normal” based on constantly shifting goalposts—whether they work and produce according to conventional measures, whether they can maintain the social order in a profoundly racist and classist society. People with disabilities often feel inferior to people without disabilities, experience guilt when requiring assistance, and find it hard to believe they are entitled to accommodations, access, education, employment, parenthood, and a host of experiences that non-disabled persons take for granted. This “burden complex” is an emotional cost exacted by internalized ableism. [16,17]

There is an unspoken requirement that people with disabilities must prove their competence to people without disabilities, must inspire, and must accept high rates of underemployment or unemployment and poverty. In 2023 the employment rate was 37.1% for people with disabilities ages 16 to 64, compared with 75% for people the same age without disabilities [18]. Employment creates its own problems, though, as few distinguish between disability and impairment. “The ADA covers technology or adaptations to allow people with disabilities to do their jobs provided it doesn’t take us longer,” says Bonneau, “and that’s the rub. If a person with disabilities confesses in a job interview that she may require extra time to meet arbitrary deadlines, will she be hired?” Bonneau avoids this problem through self-employment.

While disabled persons should individualize their own paths to functioning and flourishing, the medical community should also approach care more broadly. When clinicians try to understand a patient’s unique challenges and circumstances, asking questions through a lens of symptoms, strengths, and environmental factors instead of diagnosis may be useful. When Daltroy et al [19] developed PODCI, they worked beautifully with patients and families to do exactly this, aiming to address disorders with musculoskeletal impact in a purposely transdiagnostic design. Though reviewers typically term PODCI a “generic outcomes measure” and contrast it with disease-specific measures, it is *not* generic like PROMIS. The “precision medicine” approach focuses narrowly on the ICF’s Body Structure and Function domain, and while we do not aim to denigrate disease-specific measures, we do wish to highlight the benefit of a broader transdiagnostic approach. Our work demonstrates the need for both disease-specific assessments and treatments as well as transdiagnostic assessments and treatments. PODCI is a significant transdiagnostic tool because it helps clinicians focus on symptoms that are common across diagnoses and that affect patients’ everyday lives (two other articles are in preparation, one comparing CP population with general population and one evaluating contextual factors).

The transdiagnostic approach is a method of finding commonalities in functioning and well-being and of finding management tools to address those commonalities. As a group of disorders affecting motor functioning, CP therefore needs a transdiagnostic approach. PODCI has ecological validity with daily performance in this group of children with CP using SFA. PODCI has also been reported in the literature for a wide variety of diagnoses with musculoskeletal impact, including Normative, CP (most commonly), Brachial Plexus Birth Palsy, and Unilateral Congenital Below Elbow Deficiency, extending to obesity, fractures, sports injuries, tumors, etc. Because PODCI has met its original goals and has been used in research and outcomes assessment, it is perfectly structured for transdiagnostic assessment in this wide variety of diagnoses with musculoskeletal impact and in CP specifically. Also, PODCI is public domain; PedsQL is proprietary.

## CONCLUSIONS

The ICF framework reminds clinicians to take a broader consideration of health conditions impacting people’s lives. It encourages use of the transdiagnostic approach, paying attention to community functioning and well-being. PODCI has demonstrated its strength in accurately portraying performance in the lives of school children with CP. It has been used successfully across many diagnoses, and it has yielded a normative database (reported in the comparison article of CP to General Population) not available with disease-specific measures. The impact of CP upon individuals’ lives extends far beyond the narrow focus of their diagnosis and is interwoven with our society. We cannot neglect the broader, more inclusive focus that the transdiagnostic approach affords.

### Shortcomings

The multicenter study includes only ambulatory children with CP. Proxy responses are used for the children, and the authors acknowledge that additional information could be obtained in gathering patient responses. Some measures used (PedsQL and GMFM) had a limited number of assessments, which may affect correlations and effect sizes; for this reason, clinic measures were not compared against one another.

## Supporting information

PODCI Baseline Child 2-18 Parent Form

## Data Availability

Data is present but not available to be published.

