## Supplementary material for "Transdiagnostic Approach in Cerebral Palsy": PODCI Baseline Child 2-18 Parent Form

**(BASELINE)**

### TO BE COMPLETED BY THE PARENTS OF CHILDREN 2-18 YEARS OLD

We are asking you to complete this questionnaire about your child to better understand his/her health in general and problems related to bone and muscle conditions. Your completion of this questionnaire is voluntary. Your responses will be held in the strictest of confidence. It will take about 15 to 20 minutes to complete.

Please answer every question. Some questions may look like others, but each one is different.

Answer the questions by circling the appropriate number or by writing the answer as requested.

There are no right or wrong answers. If you are not sure how to answer a question, please give the best answer you can and make a comment in the margin. We will read all your comments, so feel free to make as many as you wish.

Return to: **Shriners Hospitals for Children – Shreveport**

1. Your Child's Name: ____________________

2. Today's Date: ____________________

3. Your Child's Birth Date: ____________________

**OFFICE USE ONLY**

Shriners Patient ID:

APPENDIX B

**Be sure to fill out both the front and back of all subsequent pages.**

**OFFICE USE ONLY**

Notes by clinician administering questionnaire:

For your child's **right side** please indicate those areas that bother your child or limit his or her function.

For your child's **left side** please indicate those areas that bother your child or limit his or her function.

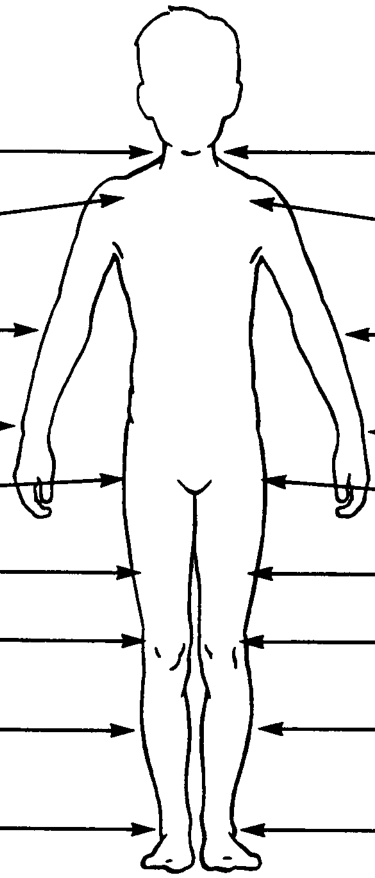


---

For your child's **back** please indicate those areas that bother your child or limit his or her function.

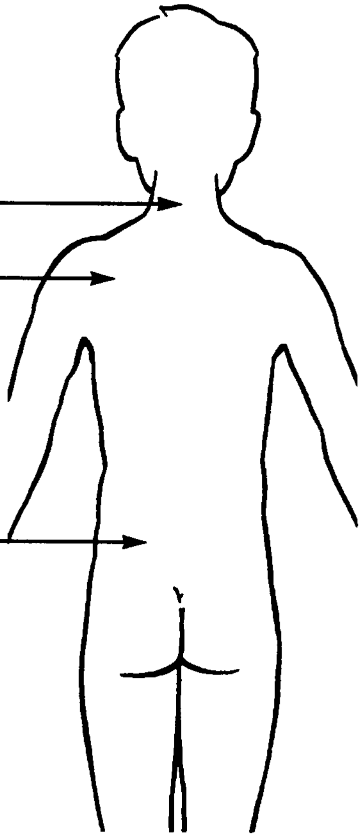

| | Excellent | Very Good | Good | Fair | Poor |
|---|---|---|---|---|---|
| 5. In general, would you say your child's health is: (Circle one number) | 1 | 2 | 3 | 4 | 5 |

| | Much better than one year ago | Somewhat better than one year ago | About the same | Somewhat worse than one year ago | Much worse than one year ago |
|---|---|---|---|---|---|
| 6. **Compared to one year ago**, how would you rate your child's health in general now? (Circle one number) | 1 | 2 | 3 | 4 | 5 |

Have you ever been told by a doctor, nurse, teacher, or other health professional that your child has had any of the following conditions? (Please circle "yes" for all conditions that apply). If yes, indicate if your child is being treated for this condition and if your child is limited by those conditions.

| | Has your child ever had it? | Does your child receive treatment for it now? | Are your child's activities limited by it now? |
|---|---|---|---|
| 7. Juvenile arthritis (one or two joints). | Yes No | Yes No | Yes No |
| 8. Juvenile arthritis (many joints). | Yes No | Yes No | Yes No |
| 9. Anorexia or bulimia (eating disorders). | Yes No | Yes No | Yes No |
| 10. Asthma. | Yes No | Yes No | Yes No |
| 11. Attention or behavioral problems. | Yes No | Yes No | Yes No |
| 12. Chronic allergies or sinus trouble. | Yes No | Yes No | Yes No |
| 13. Developmental delay. | Yes No | Yes No | Yes No |
| 14. Mental retardation. | Yes No | Yes No | Yes No |
| 15. Diabetes. | Yes No | Yes No | Yes No |
| 16. Epilepsy (seizure disorder). | Yes No | Yes No | Yes No |
| 17. Hearing impairment or deafness. | Yes No | Yes No | Yes No |
| 18. Heart problem. | Yes No | Yes No | Yes No |
| 19. Learning problem. | Yes No | Yes No | Yes No |
| 20. Sleep disturbance. | Yes No | Yes No | Yes No |
| 21. Speech problems. | Yes No | Yes No | Yes No |
| 22. Vision problems. | Yes No | Yes No | Yes No |

Some kinds of problems can make it hard to do many activities, such as eating, bathing, school work and playing with friends. We would like to find out how your child is doing.

During the **last week** was it easy or hard for your child to:

| | Easy | A little hard | Very hard | Can't do at all | Too young for this activity |
|---|---|---|---|---|---|
| 23. Lift heavy books? | 1 | 2 | 3 | 4 | 5 |
| 24. Pour a half gallon of milk? | 1 | 2 | 3 | 4 | 5 |
| 25. Open a jar that has been opened before? | 1 | 2 | 3 | 4 | 5 |
| 26. Use a fork and spoon? | 1 | 2 | 3 | 4 | 5 |
| 27. Comb his/her hair? | 1 | 2 | 3 | 4 | 5 |
| 28. Button buttons? | 1 | 2 | 3 | 4 | 5 |
| 29. Put on his/her socks? | 1 | 2 | 3 | 4 | 5 |
| 30. Write with a pencil? | 1 | 2 | 3 | 4 | 5 |

| | Rarely | Once a month | Two or three times a month | Once a week | More than once a week | Does not attend school, etc. |
|---|---|---|---|---|---|---|
| 31. On average, **over the last 12 months**, how often did your child miss school (preschool, day care, camp, etc.) because of his/her health? | 1 | 2 | 3 | 4 | 5 | 6 |

During the **last week**, how happy has our child been with:

| | Very happy | Somewhat happy | Not sure | Somewhat unhappy | Very unhappy | Child is too young |
|---|---|---|---|---|---|---|
| 32. How he/she looks? | 1 | 2 | 3 | 4 | 5 | 6 |
| 33. His/her body? | 1 | 2 | 3 | 4 | 5 | 6 |
| 34. What clothes or shoes he/she can wear? | 1 | 2 | 3 | 4 | 5 | 6 |
| 35. His/her ability to do the same things his/her friends do? | 1 | 2 | 3 | 4 | 5 | 6 |
| 36. His/her health in general? | 1 | 2 | 3 | 4 | 5 | 6 |

During the **last week**, how much of the time:

| | Most of the time | Some of the time | A little of the time | None of the time |
|---|---|---|---|---|
| 37. Did your child feel sick and tired? | 1 | 2 | 3 | 4 |
| 38. Was your child full of pep and energy? | 1 | 2 | 3 | 4 |
| 39. Did pain or discomfort interfere with your child's activities? | 1 | 2 | 3 | 4 |

During the **last week**, has it been easy or hard for your child to:

| | Easy | A little hard | Very hard | Can't do at all | Too young for this activity |
|---|---|---|---|---|---|
| 40. Run short distances? | 1 | 2 | 3 | 4 | 5 |
| 41. Bicycle or tricycle? | 1 | 2 | 3 | 4 | 5 |
| 42. Climb three flights of stairs? | 1 | 2 | 3 | 4 | 5 |
| 43. Climb one flight of stairs? | 1 | 2 | 3 | 4 | 5 |
| 44. Walk more than a mile? | 1 | 2 | 3 | 4 | 5 |
| 45. Walk three blocks? | 1 | 2 | 3 | 4 | 5 |
| 46. Walk one block? | 1 | 2 | 3 | 4 | 5 |
| 47. Get on and off a bus? | 1 | 2 | 3 | 4 | 5 |

| | Never | Sometimes | About half the time | Often | All the time |
|---|---|---|---|---|---|
| 48. How often does your child need help from another person for walking and climbing? | 1 | 2 | 3 | 4 | 5 |

| | Never | Sometimes | About half the time | Often | All the time |
|---|---|---|---|---|---|
| 49. How often does your child use assistive devices (such as braces, crutches, or wheelchair) for walking and climbing? | 1 | 2 | 3 | 4 | 5 |

During the **last week**, has it been easy or hard for your child to:

| | Easy | A little hard | Very hard | Can't do at all | Too young for this activity |
|---|---|---|---|---|---|
| 50. Stand while washing his/ her hands and face at a sink? | 1 | 2 | 3 | 4 | 5 |
| 51. Sit in a regular chair without holding on? | 1 | 2 | 3 | 4 | 5 |
| 52. Get on and off a toilet or chair? | 1 | 2 | 3 | 4 | 5 |
| 53. Get in and out of bed? | 1 | 2 | 3 | 4 | 5 |
| 54. Turn door knobs? | 1 | 2 | 3 | 4 | 5 |
| 55. Bend over from a standing position and pick up something off the floor? | 1 | 2 | 3 | 4 | 5 |

| | Never | Sometimes | About half the time | Often | All the time |
|---|---|---|---|---|---|
| 56. How often does your child need help from another person for sitting and standing? | 1 | 2 | 3 | 4 | 5 |
| 57. How often does your child use assistive devices (such as braces, crutches, or wheelchair) for sitting and standing? | 1 | 2 | 3 | 4 | 5 |

| | Yes, easily | Yes, but a little hard | Yes, but very hard | No |
|---|---|---|---|---|
| 58. Can your child participate in **recreational outdoor activities** with other children the same age? (For example: bicycling, tricycling, skating, hiking, jogging) | 1 | 2 | 3 | 4 |

**If you answered "no" to Question 58 above**, was your child's activity limited by: (Circle "yes" to all that apply.)

| | Yes |
|---|---|
| 59. Pain? | 1 |
| 60. General Health? | 1 |
| 61. Doctor or parent instructions? | 1 |
| 62. Fear the other kids won't like him/ her? | 1 |
| 63. Dislike of recreational outdoor activities? | 1 |
| 64. Too young? | 1 |
| 65. Activity not in season? | 1 |

| | Yes, easily | Yes, but a little hard | Yes, but very hard | No |
|---|---|---|---|---|
| 66. Can your child participate in **pickup games or sports** with other children the same age? (For example: tag, dodge ball, basketball, soccer, catch, jump rope, touch football, hop scotch) | 1 | 2 | 3 | 4 |

**If you answered "no" to Question 66 above**, was your child's activity limited by: (Circle "yes" to all that apply.)

| | Yes |
|---|---|
| 67. Pain? | 1 |
| 68. General Health? | 1 |
| 69. Doctor or parent instructions? | 1 |
| 70. Fear the other kids won't like him/ her? | 1 |
| 71. Dislike of pickup games or sports? | 1 |
| 72. Too young? | 1 |
| 73. Activity not in season? | 1 |

| | Yes, easily | Yes, but a little hard | Yes, but very hard | No |
|---|---|---|---|---|
| 74. Can your child participate in **competitive level sports** with other children the same age? (For example: hockey, basketball, soccer, football, baseball, swimming, running [track or cross country], gymnastics, or dance) | 1 | 2 | 3 | 4 |

**If you answered "no" to Question 74 above**, was your child's activity limited by: (Circle "yes" to all that apply.)

| | Yes |
|---|---|
| 75. Pain? | 1 |
| 76. General Health? | 1 |
| 77. Doctor or parent instructions? | 1 |
| 78. Fear the other kids won't like him/ her? | 1 |
| 79. Dislike of competitive level sports? | 1 |
| 80. Too young? | 1 |
| 81. Activity not in season? | 1 |

| | Often | Sometimes | Never or rarely |
|---|---|---|---|
| 82. How often in the **past week** did your child get together and do things with friends? | 1 | 2 | 3 |

**If you answered "sometimes" or "never or rarely" to Question 82 above**, was your child's activity limited by: (Circle "yes" to all that apply.)

| | Yes |
|---|---|
| 83. Pain? | 1 |
| 84. General health? | 1 |
| 85. Doctor or parent instructions? | 1 |
| 86. Fear the other kids won't like him/her? | 1 |
| 87. Friends not around? | 1 |

| | Often | Sometimes | Never or rarely | No gym or recess |
|---|---|---|---|---|
| 88. How often in the **past week** did your child participate in gym/ recess? | 1 | 2 | 3 | 4 |

**If you answered "sometimes" or "never or rarely" to Question 88 above**, was your child's activity limited by: (Circle "yes" to all that apply.)

| | Yes |
|---|---|
| 89. Pain? | 1 |
| 90. General health? | 1 |
| 91. Doctor or parent instructions? | 1 |
| 92. Fear the other kids won't like him/her? | 1 |
| 93. Dislike of gym/recess? | 1 |
| 94. School not in session? | 1 |
| 95. Does not attend school? | 1 |

| | Usually easy | Sometimes easy | Sometimes hard | Usually hard |
|---|---|---|---|---|
| 96. Is it easy or hard for your child to make friends with children his/ her own age? | 1 | 2 | 3 | 4 |

| | None | Very mild | Mild | Moderate | Severe | Very severe |
|---|---|---|---|---|---|---|
| 97. How much pain has your child had during the **last week**? | 1 | 2 | 3 | 4 | 5 | 6 |

| | Not at all | A little bit | Moderately | Quite a bit | Extremely |
|---|---|---|---|---|---|
| 98. During the last week, how much did pain interfere with your child's normal activities (including at home, outside of the home, and at school)? | 1 | 2 | 3 | 4 | 5 |

What expectations do you have for your child's treatment?
As a result of my child's treatment, I expect my child:

| | **Definitely yes** | **Probably yes** | **Not sure** | **Probably not** | **Definitely not** |
|---|---|---|---|---|---|
| 99. To have pain relief. | 1 | 2 | 3 | 4 | 5 |
| 100. To look better. | 1 | 2 | 3 | 4 | 5 |
| 101. To feel better about himself/ herself. | 1 | 2 | 3 | 4 | 5 |
| 102. To sleep more comfortably. | 1 | 2 | 3 | 4 | 5 |
| 103. To be able to do activities at home. | 1 | 2 | 3 | 4 | 5 |
| 104. To be able to do more at school. | 1 | 2 | 3 | 4 | 5 |
| 105. To be able to do more play or recreational activities (biking, walking, doing things with friends). | 1 | 2 | 3 | 4 | 5 |
| 106. To be able to do more sports. | 1 | 2 | 3 | 4 | 5 |
| 107. To be free from pain or disability as an adult. | 1 | 2 | 3 | 4 | 5 |

| | **Very satisfied** | **Somewhat satisfied** | **Neutral** | **Somewhat dissatisfied** | **Very dissatisfied** |
|---|---|---|---|---|---|
| 108. If your child had to spend the rest of his/ her life with his/ her bone and muscle condition **as it is right now**, how would you feel about it? | 1 | 2 | 3 | 4 | 5 |

109. What is your child's gender?

❑ Male ❑ Female

110. What is your child's race? (check all that apply)

❑ White ❑ Black or African-American ❑ Hispanic
❑ Asian or Pacific Islander ❑ Native American Indian ❑ Other (please specify) ________________________

111. Who lives at home with your child? (check all that apply)

❑ Mother ❑ Stepmother ❑ Foster mother ❑ Father ❑ Stepfather
❑ Foster father ❑ Brothers and/or sisters (How many _______ ?) ❑ Other adults

112. What is your relationship to this child?

❑ Mother ❑ Stepmother ❑ Foster mother ❑ Father ❑ Stepfather
❑ Foster father ❑ Brother ❑ Sister ❑ Grandmother ❑ Grandfather
❑ Aunt ❑ Uncle ❑ Guardian ❑ Other ________________________

Please answer the next two questions about **your health (not your child's)**:

113. In general, would you say your health is:

❑ Excellent ❑ Very good ❑ Good ❑ Fair ❑ Poor

---

114. Compared to **one year ago**, how would you rate **your health** in general now?

❑ Much better now than 1 year ago

❑ Somewhat better now than 1 year ago

❑ About the same as 1 year ago

❑ Somewhat worse now than 1 year ago

❑ Much worse now than 1 year ago
